# Antibiotic Treatment Duration for Bacterial Infections - A Systematic Review and Critical Appraisal

**DOI:** 10.1101/2022.12.11.22283311

**Authors:** Yin Mo, Wei Cong Tan, Ben S. Cooper

## Abstract

Reducing antibiotic treatment duration is a key stewardship intervention to mitigate antimicrobial resistance (AMR). We performed a systematic review of antibiotic duration randomised controlled trials for treatment or prevention of bacterial infections in humans, appraised their methodologies and identified gaps in evidence. All randomised trials indexed in MEDLINE and EMBASE up to October 2022 which compared different treatment durations were included. We found 296 trials, of which 85% showed equivalence or non-inferiority. The majority (73%) considered treatment for respiratory tract infections, genitourinary infections, and post-surgical prophylaxis. Few trials studied severe infections, such as bloodstream infections and ventilator-associated pneumonia. Trial designs shifted from superiority (74%, 178/242 before 2016) to non-inferiority (74%, 40/54 after 2016). 94% of the trials that defined a per-protocol population reported non-adherence, which may increase the probability of concluding non-inferiority. Only 9 trials collected data to demonstrate the effect of antibiotic duration on AMR colonisation.

**Registration:** PROSPERO 2021 CRD42021276209

**Funding:** National Research Foundation Singapore Central Gap Fund; National Medical Research Council, Singapore; Medical Research Council, UK

## Introduction

Antimicrobial resistance (AMR) represents a global public health crisis. Human consumption of antibiotics is a major driver of AMR, [1] and reducing unnecessary antibiotic use is a key intervention for reducing AMR. Increasing calls to view antibiotics as a non-renewable resource gave rise to systematic and evidence-based antibiotic stewardship programmes in the early 2000s. [2] These programmes are strongly advocated by the World Health Organization (WHO) and international infectious disease authorities.

The key antibiotic stewardship strategies can be broadly categorised as i) starting antibiotic treatments only when a bacterial infection is clearly indicated, ii) administering the optimal choice and dosing of antibiotic by considering the host, site of infection, pathogen, and pharmacokinetic/ pharmacodynamic properties of the drug, and iii) limiting duration of treatment. [3–5] Reducing antibiotic treatment duration is one of the most commonly implemented antibiotic stewardship strategies reported in the literature, [6] and the one that is deemed to be safest and most acceptable by practising clinicians. [7] This includes shortening courses for established bacterial infections, preventing bacterial infections, as well as rapid discontinuation of prescriptions after bacterial infections are ruled out.

Prior to heightened concern about AMR, the conventional principle for duration of antibiotic therapy was to treat beyond clinical improvement in order to prevent both relapse of infection and development of antibiotic resistance. The message of always completing antibiotic courses to prevent the development of resistance has remained widespread until recent years, promoted by the WHO, international health authorities, national health campaigns and school curricula. [7, 8]

Early knowledge about treatment failures with inadequate antibiotic duration emerged in the 1940s when penicillin was first used in clinical medicine. Abraham *et al*. reported in detail a case series involving patients with staphylococcal and streptococcal sepsis. [9] This was during a time when penicillin was produced in limited amounts locally in the laboratory and repeated doses of the drug were recovered from the patients’ urine. Patients who initially responded to treatment eventually succumbed due to insufficient availability of drugs.

When mass production of penicillin became available in the mid-1940s, Keefer *et al*. found that the majority of 500 patients with community-acquired pneumonia recovered with only 2 to 3 days of penicillin. [10] In the following years, Dawson, Tillett, Meads *et al.* confirmed these findings, [11–13] but reported two instances of relapses. [13] They also identified subgroups of patients who might require longer treatment duration, who were primarily those with pockets of pus such as empyema and local or systemic immunocompromised illnesses such as chronic obstructive pulmonary disease. [11] These potentially life-threatening treatment failures, coupled with the observation that antibiotic side effects were rare, motivated recommendations that treatment duration for community-acquired pneumonia be 10 to 14 days until the early 2000s. [14–16]

Concerns about the development of antibiotic resistance with inadequate treatment were voiced initially by Fleming as he observed that bacteria exposed to antibiotics *in vitro* became increasingly resistant. [17] The spotlight on excessive antibiotic use driving AMR in the late 1990s sparked numerous randomised controlled trials to shorten treatment duration. Aside from directly comparing treatment durations, inflammatory markers such as C-reactive protein and procalcitonin, and individualised expert opinion such as infectious disease specialist consultations have also been used in trials to inform treatment duration decisions. [18]

This review focuses on optimising antibiotic treatment duration as an intervention to reduce AMR, one of the most practical and fundamental questions of antibiotic use. We performed a systematic review of all antibiotic duration randomised controlled trials to: i) review antibiotic treatment durations for the various bacterial infections; ii) identify gaps in evidence for antibiotic treatment durations in terms of settings, patient populations and infectious conditions; and iii) appraise trial methodologies and identify areas for improvement.

## Methods

### Search Strategy and Selection Criteria

The systematic review was performed in accordance with the Preferred Reporting Items for Systematic Reviews and Meta-Analysis (PRISMA) guidelines. [19] Randomised controlled trials published to 4 October 2022, and indexed in MEDLINE and EMBASE were sought. [20] Unpublished studies and pre-prints were excluded. This systematic review was registered on PROSPERO (reference: PROSPERO 2021 CRD42021276209).

The specific inclusion criteria were:

i. Participants: Patients from both hospital and community settings who received antibiotics for the prevention or treatment of bacterial infections.
ii. Intervention: Varying duration of antibiotic treatment. The specific interventions which were used to guide treatment duration may include the use of inflammatory markers, expert opinion or electronic prescription systems.
iii. Comparison: Patients who received antibiotic(s) of different durations.
iv. Primary or secondary outcome(s):

a. Clinical cure: Number of patients who had clinical or microbiological cure, or infection relapses, or treatment failure.
b. Colonisation: Number of patients who were colonised with antibiotic-resistant bacteria before and after antibiotic treatment. The sites of colonisation may include, but are not limited to, the digestive tract, the respiratory tract, and the urinary tract.

The exclusion criteria were:

i. Participants: Patients with non-infectious conditions; fungal or viral infections; tuberculosis.
ii. Comparison: Patients who received no antibiotic treatment, e.g. comparison between antibiotic treatment and a surgical procedure; patients who received a similar antibiotic in a slow-release formulation.
iii. Outcome: Feasibility for study intervention such as in pilot studies.

### Data Extraction and Analysis

A Boolean search strategy with search terms pertaining to antibiotic treatment, duration, bacteria infections, human health, and randomised controlled trials was adopted (Supplementary material 1). Additional relevant titles found in other systematic reviews were also included.

After removing duplicates, the identified studies were screened according to the above inclusion and exclusion criteria. Full text for all studies included in the systematic review were retrieved for data extraction. Data extracted from all included studies were:

i. year of publication;
ii. type of bacterial infection including pathogen and site of infection;
iii. treatment durations compared;
iv. blinding of participants, prescribing physicians and investigators;
v. intervention to vary treatment duration;
vi. healthcare setting;
vii. age group of participants; and
viii. country/countries where patients were enrolled from.

To provide a more detailed assessment of current methodologies and aspects of trial design and conduct affecting risk of bias, additional data were extracted from trials conducted in the last 15 years (from 2006 onwards). These additional data extracted included:

i. study hypothesis and sample size calculation;
ii. how the bacterial infection was identified as described in the inclusion criteria;
iii. choice of antibiotic treatment;
iv. randomisation process;
v. follow-up period;
vi. funding;
vii. non-adherence and how this was monitored;
viii. analysis methods;
ix. primary outcome;
x. whether antibiotic side effects and health-economic outcomes were reported.

Randomised trials published from 2006 onwards, and which achieved the target enrolment number, were further assessed according to the key domains outlined in the revised tool for assessing risk of bias in randomised trials (RoB 2). [21] These domains included risk of bias arising from the randomisation process, deviations from the intended interventions, measurement and quality of outcome data, and selection of the reported result. In addition, quality of the statistical analysis was assessed by considering sample size calculations, choice of non-inferiority or equivalence margins and appropriateness in the choice of outcome.

## Results

### Overview of antibiotic treatment duration randomised trials

The initial electronic database search produced 2649 unique records published from 1969 to October, 2022 (Figure 1). Out of these, 296 fulfilled the inclusion criteria. 85% (253/296) of the trials concluded that there was either no statistical difference, equivalence or non-inferiority between the short and long treatment durations when clinical outcomes were compared. The median number of study participants randomised in the trials was 229 (IQR 100 to 435). 94 (32%) of the trials were double-blinded, where placebo was administered for those randomised to the short duration to complete antibiotics courses. These were trials that usually administered only one or two types of antibiotic treatment to the participants.

**Figure 1:**
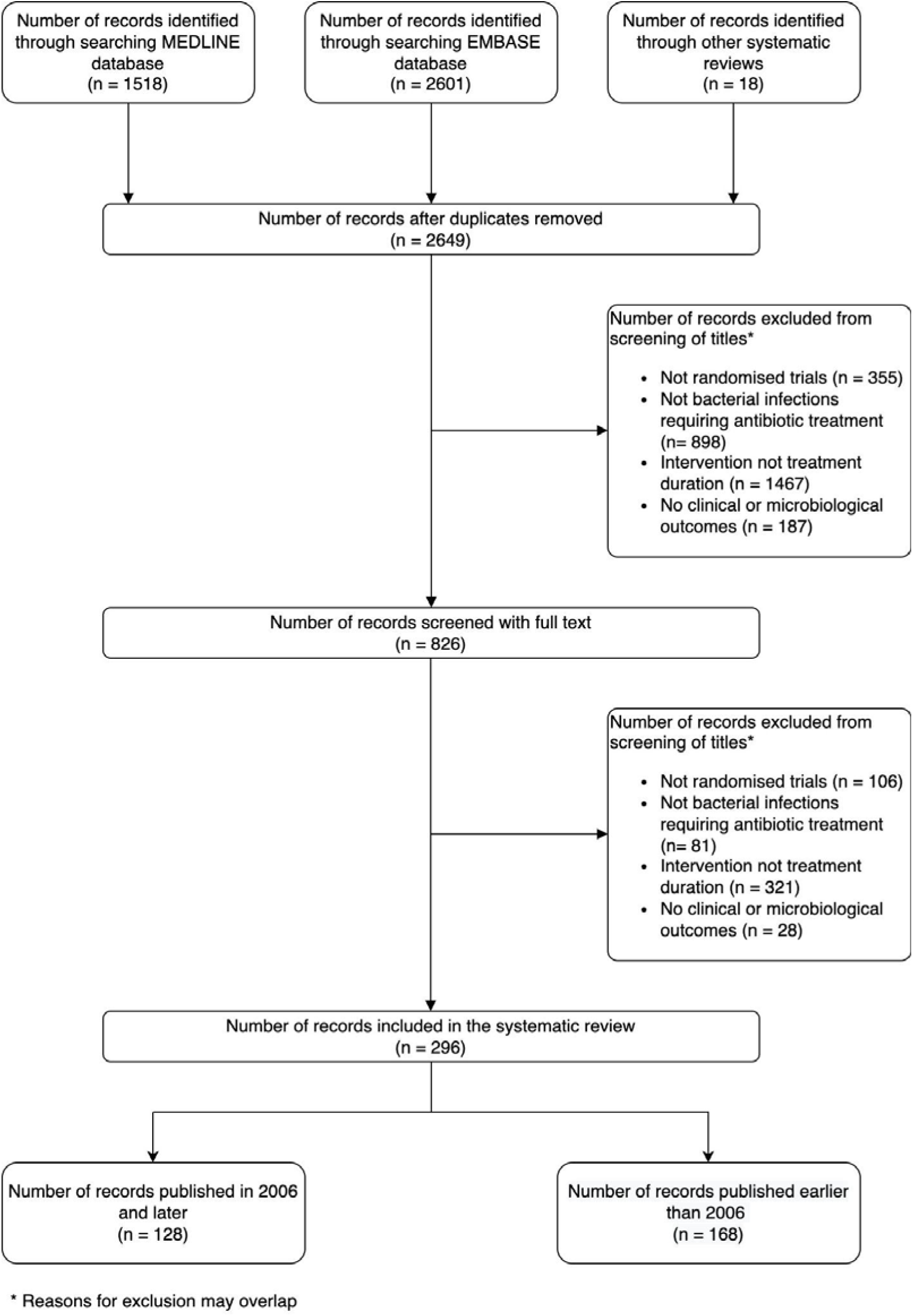
PRISMA flow diagram for systematic review on antibiotic duration randomised trials.

The most frequently studied bacterial infections were upper (53/296, 18%) and lower respiratory tract infections (64/296, 22%), genitourinary infections (49/296, 17%), and post-surgical prophylaxis (49/296, 17%) (Figure 2). 21 trials (7%) studied conditions with an infection focus that required surgical removal or debridement, and in 14 of these trials (14/21, 67%) the protocol mandated satisfactory source control in addition to antibiotic treatment.

**Figure 2:**
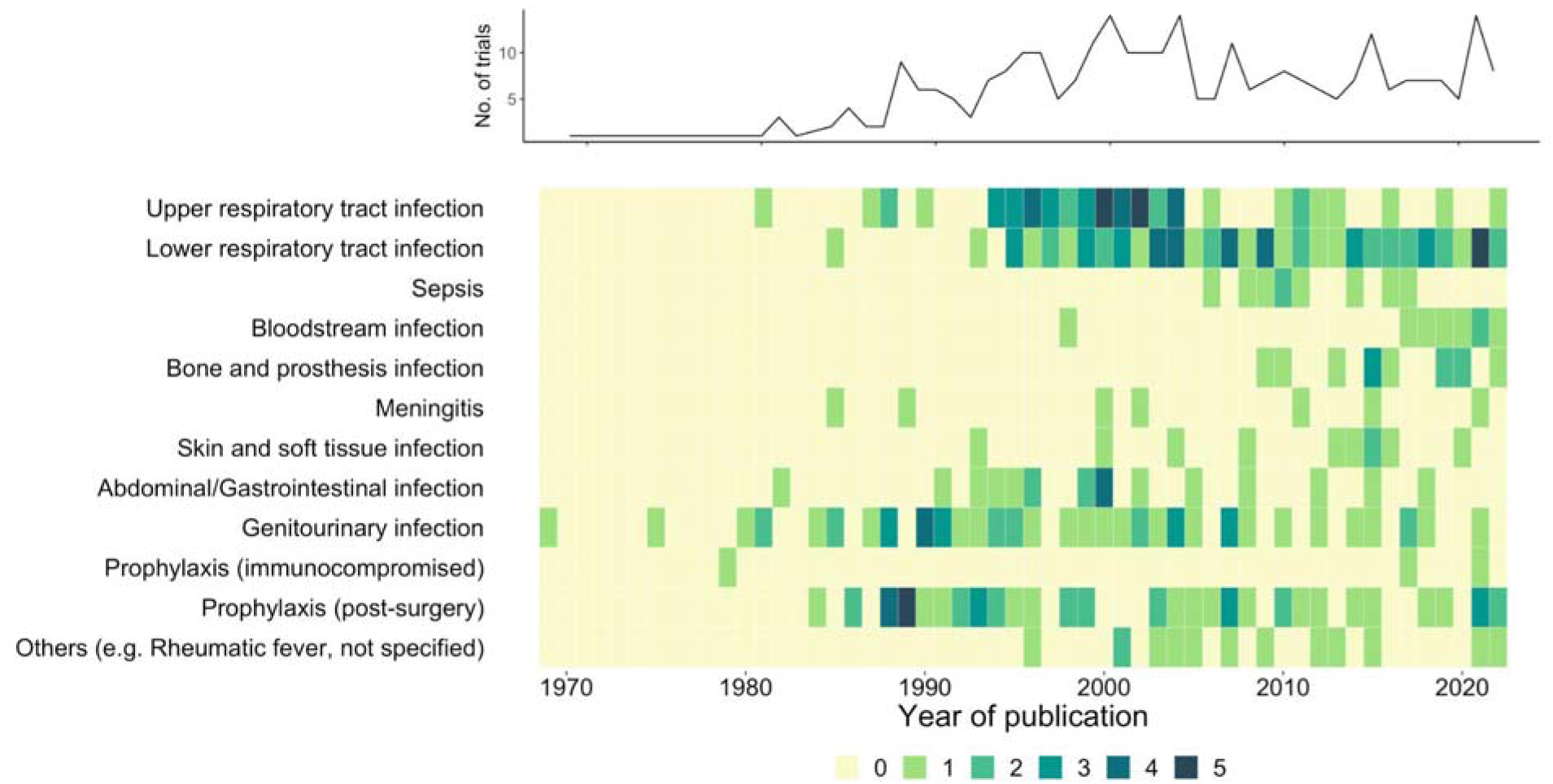
Types of bacterial infections studied by antibiotic duration randomised trials over time. The top panel shows the total number of published randomised trials per year from 1969 to October, 2022. The bottom panel shows the number of trials for each type of infection over time. The shading intensity of the green squares represents the number of trials per year.

In terms of patient characteristics, the most frequently represented groups consisted of adults from either the hospital general ward or the community in upper- and upper-middle income countries (Figure 3). Only 23 (8%) of the trials were conducted in intensive care settings, and 11 trials (4%) enrolled patients from lower-middle or low income countries.

**Figure 3:**
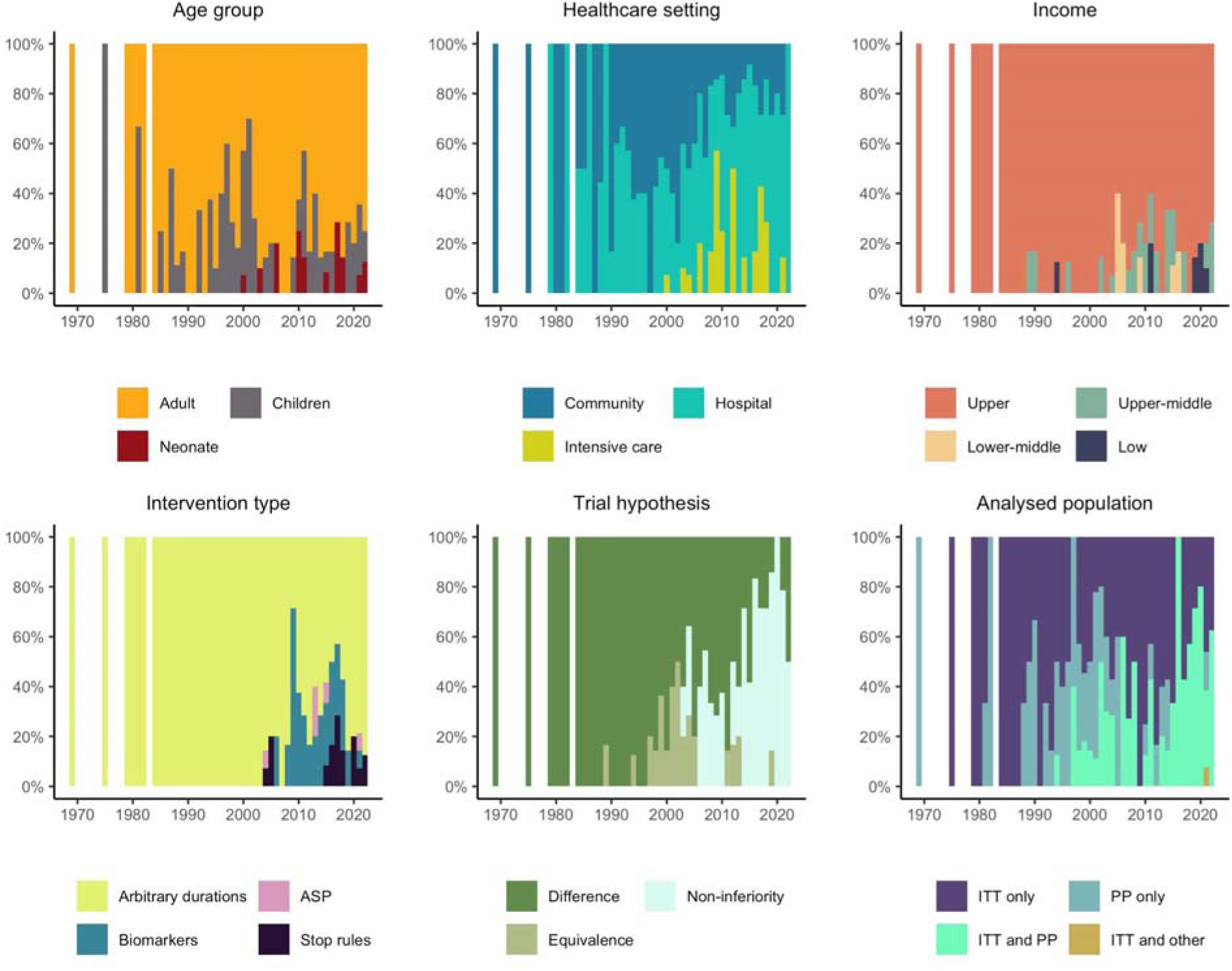
Characteristics of antibiotic duration trials over time. Each panel is labelled with a trial characteristic. These characteristics include i) age group of the trial participants, ii) the healthcare setting which the participants were enrolled from, iii) the minimum income level of the country/countries where the participants were enrolled from, iv) the type of intervention studied in the trials, v) the trial hypothesis design, and vi) the participant populations which the final conclusion of the trials were based on. ITT: intention-to-treat; PP: per protocol; ASP: antibiotic stewardship. Proportion of the trials published each year with a certain characteristic (y-axis) is plotted against the year of publication (x-axis) to illustrate the changes in these trial characteristics over time.

Trial designs and methodologies have evolved with time (Figure 3). Prior to 2004, the antibiotic treatment duration trials exclusively tested various arbitrary durations. Starting in the late 2000s, biomarkers such as procalcitonin and C-reactive proteins were increasingly studied, but there has been a decrease in such studies since the mid-2010s. Another type of intervention that has emerged in recent years is the use of treatment protocols incorporating antibiotic stopping rules to terminate treatment according to individual clinical response.

Trial designs have shifted from superiority, which accounted for 74% (178/242) of trials prior to 2016, to non-inferiority for trials published after 2016 (40/54, 74%). The study populations analysed for the trial hypothesis have also changed from only the intention-to-treat group (135/239 trials, 56%) prior to 2016 to also including the per-protocol participants (31/53 trials, 59%) after 2016. The intention-to-treat analysis compares outcomes in groups defined by the initial random allocation regardless of the actual treatment received. The per-protocol analysis includes only those patients who received the originally allocated treatment.

### Quality of trial design, conduct and analysis

#### • Intervention

Of 128 trials published in the last 15 years, 65 (51%) included multiple antibiotic regimens within the randomisation arms. These included all of the trials using biomarkers and antimicrobial stewardship as the intervention. An additional 21 trials (16%) compared different antibiotics between the short and long arms, such as 5-day nitrofurantoin vs single-dose fosfomycin for the treatment of uncomplicated urinary tract infection. [22] In 57 trials (45%), the antibiotics prescribed were culture-directed, while the rest were empirical.

23 trials (18%) were funded by a pharmaceutical or diagnostics company. Amongst these industry-funded trials, 5 studied a biomarker as the intervention while all others studied a particular type of antibiotic sponsored by the company.

#### • Adherence to intervention and study population analysed

The sample sizes and analysed participant populations are shown in (Figure 4). In 70 trials (70/128, 55%), some randomised patients were left out of the final analysis (median 4%, IQR 2 to 12%). The commonest reasons for exclusion of these participants were protocol deviations, loss to follow-up and consent withdrawal.

**Figure 4.**
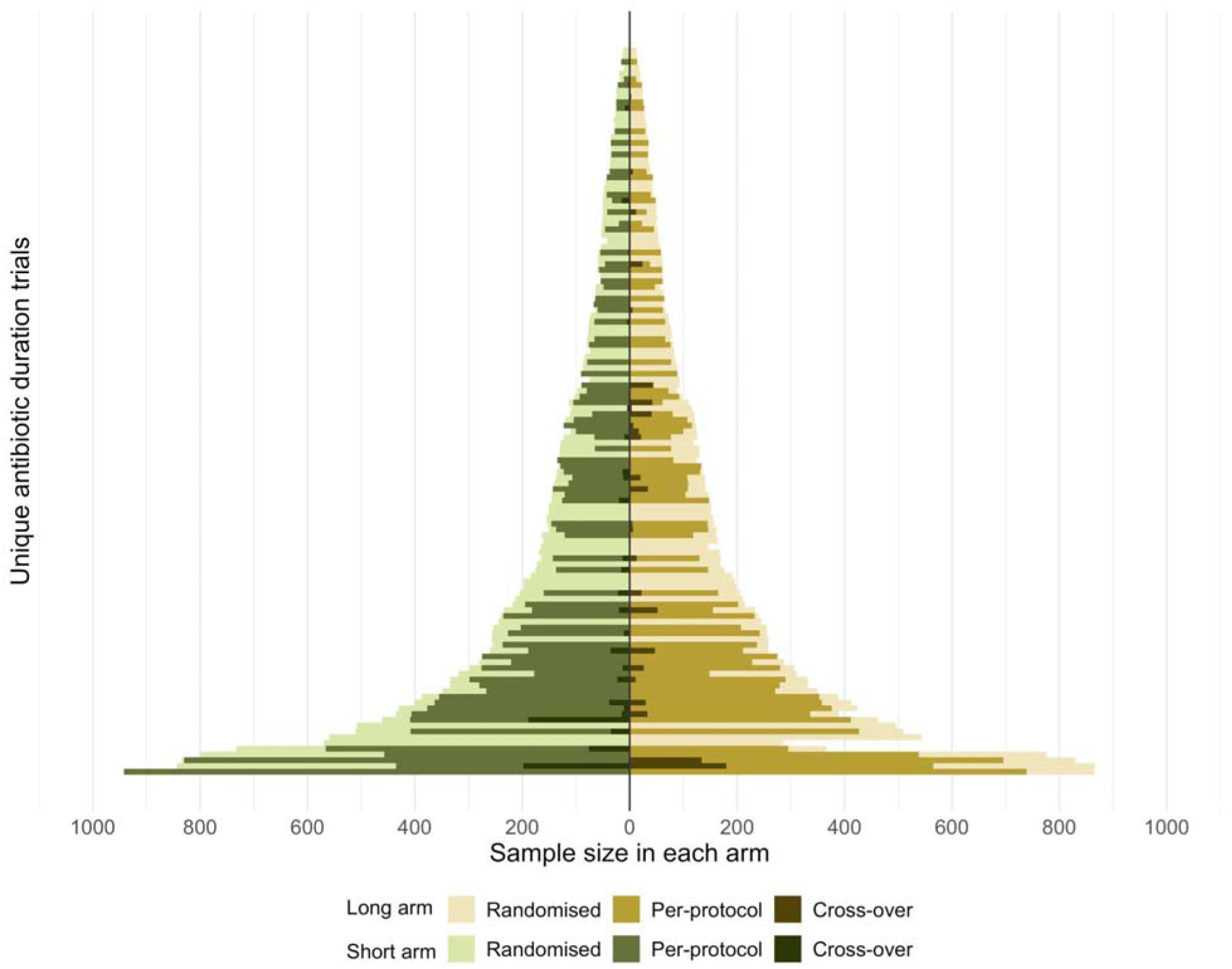
Comparison of randomised, analysed and per-protocol trial participants in antibiotic duration trials 2006-2021. One hundred and four antibiotic duration randomised trials are presented in the graph. Each horizontal bar represents a unique trial. One trial published within the period 2006-2021 with a sample size of more than 3000 participants is not shown on the graph to ensure visibility of the bars plotted from other trials. The green bars on the left represent trial participants who were randomised to the long duration arm; the gold bars on the right represent participants randomised to the short duration arm. The shade of the colours represents different populations of trial participants: the widest bars on each row represent the number of participants randomised in each trial (lightest colours); followed by the number of participants who were reported to be per-protocol (darker shades); the darkest shades represent the number of participants who crossed over to the opposite arm, i.e. participants allocated to the short duration arm who were actually treated with antibiotics for a longer duration and vice versa.

Adherence to randomised intervention was reported in terms of either the proportion of participants who consumed antibiotics for a certain number of days (frequently a criterion used to define the per-protocol population, i.e. at or above a threshold number of days as adherence for long arm, while at or below a second lower threshold as adherence for short arm) or as the mean antibiotic duration actually observed in the respective randomisation arms (i.e. adherence was not reported according to a pre-specified protocol but by observed values). 94 trials (73%) reported proportions of participants who were adherent, while 49 trials (38%) reported mean antibiotic durations observed in either arms. Just over half of those trials that reported a measure for adherence (50/96, 52%) reported participants from the short arm crossing over to the long arm (median 3%, IQR 0 to 14%). On the other hand, 49 trials (49/96, 51%) reported participants from the long arm crossing over to the short arm (median 4%, IQR 0 to 9%).

94% of the trials that reported the per-protocol number of participants had non-adherence (73/78). The median proportion of non-adherent participants was 13% (IQR 7 to 23%). Amongst the 49 trials which reported the actual treatment durations observed in the respective randomisation arms, the difference between the actual and the intended treatment durations stated in the protocol ranged from -4 to 2 days (mean of 0 days). The commonest methods for adherence monitoring was medical chart review (46/78 trials, 59%) or self-reporting (31/78 trials, 40%).

#### • Outcome assessment

109 trials (95%) specified a follow-up period for the primary outcome, 7 of which were during hospitalisation or at the end of treatment. The median follow-up period was 28 days (IQR 15 to 40 days).

114 trials (89%) specified a primary outcome. Most used clinical recovery, infection recurrence, microbiological eradication or mortality as the primary outcome (125/128, 98%), while the others used antibiotic duration or length of hospital stay (Table 1). 96 trials (75%) included at least one primary outcome which was subjective, which most frequently involved resolution of symptoms that were self-reported or determined by clinicians. Only in 25 trials (20%) were the assessors determining the outcome blinded to the randomisation arm. 12 trials (9%) used a composite outcome.

**Table 1.** Summary of main findings from antibiotic duration randomised trials.

| Indication for antibiotic | Total number of trials | Most frequently used primary outcome | Current evidence based on randomised controlled trials | Remaining gaps in evidence/ challenges in performing randomised controlled trials |
| --- | --- | --- | --- | --- |
| **Respiratory tract** |  |  |  |  |
| Acute group A Streptococcus (GAS) pharyngotonsillitis | 25 | Resolution of acute symptoms; recurrence<br>Microbiological eradication | 5 to 7 days penicillin V adequate for symptom resolution but led to lower eradication rate than standard 10-day course [23]<br>Alternatives to penicillins include macrolides and late-generation cephalosporins, which can be used in shorter courses (3 days azithromycin approved in Europe, 5 days cefpodoxime approved by FDA) | Short-course is comparable with long-course antibiotics in terms of symptomatic resolution. [23] However, long-course penicillin or amoxicillin is the current standard due to potentially severe immune sequelae from GAS persistence in oropharynx. [24] Few trials evaluated such long-term complications from reduced antibiotic treatment duration<br>No trial evaluated the effect of an alternative antibiotic to penicillin for preventing acute rheumatic fever<br>Few trials were performed in low-to-middle income setting where prevalence of rheumatic fever remains high [25] |
| Otitis media in children | 15 | Resolution of acute symptoms; recurrence | 10 days for children <2 years or of any age with tympanic membrane perforation or history of recurrent acute otitis media [26] | Most trials were unblinded, did not use strict diagnostic criteria, compared different antibiotics in the comparison arms, and had heterogenous outcome measures to indicate clinical resolution |
|  |  |  | 5 to 7 days for older children without tympanic membrane perforation or history of recurrent acute otitis media [26] | of symptoms [27]<br>Lack of trials in older children above 2 years old, in low-to-middle income countries where prevalence of antibiotic resistance may be higher<br>High rates of spontaneous resolution suggest that many children do not need to be treated with antibiotics. However, there is a lack of trials which identified this patient subset. |
| Acute bacterial sinusitis | 14 | Resolution of acute symptoms; recurrences | 5 days in adults resulted in comparable outcomes with longer courses in terms of symptom resolution, and is associated with fewer side effects [28]<br>10 days in children recommended based on expert opinion [29] | Trials used heterogenous adjunctive therapies for symptom relief and enrolled patients with varying symptom duration prior to treatment<br>Non-specific diagnostic criteria and lack of microbiological testing may have led to patients with viral sinusitis enrolled into the trials<br>No randomised trials were performed in children |
| Community-acquired pneumonia | 31 | Resolution of acute symptoms; recurrences<br>Mortality | Antibiotic duration is catered to treatment response and severity of disease<br>Mild to moderate disease in adults: outpatient short-course antibiotics (e.g. 5 days amoxicillin, or 3 days | A wide variety of antibiotics were compared within and between trials<br>No trials compared short- versus long-course with the same antibiotic at the same daily dosage in adolescent and adult outpatients [33] |
|  |  |  | azithromycin) until resolution of fever and clinically stable [30,31]<br>Severe disease in adults: inpatient antibiotics for at least 5 days, and may be prolonged in cases with pulmonary complications or extrapulmonary dissemination, inadequate procalcitonin response, antibiotic resistance such as methicillin-resistant *S. aureus* or *Pseudomonas* spp.<br>Mild to moderate disease in children: outpatient short-course antibiotics (e.g. 7 days amoxicillin, or 5 days azithromycin) until resolution of fever and clinically stable<br>Severe disease in children: 5 to 10 days antibiotics depending on clinical response and presence of complications (based on observational data and extrapolation from adult randomised trials) [32] | Through there were a number of trials which supported 3 to 5 days antibiotics treatment in children with mild community-acquired pneumonia, lack of chest imaging and microbiological testing in these trials may have allowed patients with viral pneumonia or upper respiratory tract infections to be enrolled into trials<br>Lack of randomised controlled trials for radiologically confirmed pneumonia in children, especially in low-to-middle income settings [34–36] |
| Ventilator-associated pneumonia | 5 | Mortality; recurrence | 7 days for patients with no pulmonary or extrapulmonary complications [37,38] | Higher rate of recurrence with short-course antibiotics observed in patients with pneumonia caused by nonfermenting Gram-negative rods, such as *P. aeruginosa* [39–41] |
|  |  |  |  | Non-specific diagnostic criteria may have led to patients with no pneumonia enrolled into the trials<br>Optimal approach of using procalcitonin to stop antibiotics is not known and yet to be routinely recommended by international infectious disease and critical care societies [37,38] |
| Chronic obstructive pulmonary disease exacerbation | 20 | Symptom resolution<br>Microbiological eradication | 5 to 7 days were comparable with longer courses in terms of symptom resolution, and is associated with fewer side effects [42] | Trials adopted highly heterogeneous diagnostic criteria e.g. smoking exposure, airflow obstruction, definitions of an exacerbation, microbiological testing. This may have led to patients with bronchiectasis and chronic asthma into the trials<br>A wide variety of antibiotics were compared within and between trials |
| **Genitourinary tract** |  |  |  |  |
| Uncomplicated urinary tract infections (simple cystitis in women) | 15 | Symptom resolution<br>Microbiological eradication | Short-course (e.g. 5 days nitrofurantoin, 3 days Trimethoprim-sulfamethoxazole) | Short-course antibiotics is well established and supported by numerous randomised trials across women of different age groups |
| Complicated urinary tract infections (in males, catheter-associated, upper | 20 | Symptom resolution<br>Microbiological | 7 days, e.g. ciprofloxacin or trimethoprim-sulfamethoxazole, for males with symptomatic urinary tract infection based on a single | No trials evaluated antibiotic duration directly in men with simple cystitis and catheter-associated urinary tract infection (in the only randomised trial |
| urinary tract involvement) |  | eradication | randomised trial [43]<br>7 to 14 days depending on clinical response and antibiotic choice based on expert opinion and observational data [44,45]<br>5 to 14 days depending on clinical response and antibiotic choice (e.g 5 days fluoroquinolone) [46,47] | for catheter-associated urinary tract infection, urinary catheter was retained in the long-course arm but removed in the short-course arm) [48]<br>Acute complicated urinary tract infections are highly heterogeneous due to e.g. the presence of nephrolithiasis or foreign bodies, various sites of infection and infection severity |
| Bacterial vaginosis | 4 | Symptom resolution; recurrence<br>Vagina pH, presence of clue cells and whiff test | Short-course oral or topical antibiotics e.g. 7 days oral clindamycin or metronidazole based on expert opinion; single dose of secnidazole or 2 days of tinidazole as alternatives [49] | Incomplete understanding of disease pathology related to vaginal microbiota disturbances, associated with multiple bacterial species<br>High rate of recurrence (>50%) with current recommended treatment [50]<br>Limited number of trials evaluated optimal antibiotic duration |
| **Bone and joint infections** |  |  |  |  |
| Adult osteomyelitis/ septic arthritis and prosthetic joint infection | 7 | Mortality; clinical cure; recurrence | Typically >4 weeks depending on adequacy of surgical debridement/ prosthesis removal and response to treatment (e.g. >4 weeks for hematogenous non-vertebral osteomyelitis, 4 to 6 weeks after 2-stage exchange arthroplasty, potentially life-long suppression for retained infected implants) [51–54] | Highly heterogeneous infection, e.g. joints involved, severity/ complications of infections, time from symptom onset to surgical debridement, various bacterial pathogens, which brings challenge to defining inclusion and exclusion criteria in trials<br>Interventions applied in the trials were<br>also varied e.g. single versus 2-stage arthroplasty, removal versus retention of implants, requirement for surgical debridement, different antibiotic regimens with variable bioavailability to bony tissue [55] |
| Childhood osteomyelitis/ septic arthritis | 2 | Clinical cure; recurrence | Typically >4 weeks depending on response to treatment, e.g. inflammatory markers, based on expert opinion and observational data | Few trials evaluated antibiotic duration. [56] Of these, many were underpowered.<br>Non-specific enrolment criteria e.g. inclusion of patients with both culture positive and negative cases<br>Lack of generalisability in trials which mainly enrolled mild cases e.g. minimal complications, bacterial pathogens which responded quickly to antibiotic therapy e.g. methicillin-susceptible *Staphylococcus aureus* [57] |
| Diabetic foot | 3 | Clinical cure; recurrence | Individualised duration depending on clinical response, extent of infection, adequacy of surgical debridement, antibiotic regimen and bacterial pathogen [58,59] | Few trials evaluated antibiotic duration<br>Difficulties differentiating colonising bacteria and true pathogens in microbiological testing<br>Heterogeneous patient population which may include those with various diabetic control, underlying neuropathy and vascular stenosis, infection severity |
|  |  |  |  | Heterogeneous trial interventions which included various surgical debridement procedures and wound care regimens |
| **Bloodstream infections** |  |  |  |  |
| Gram-negative | 4 | Mortality;<br>microbiological eradication;<br>recurrence | 7 to 14 days depending on clinical response, eradication of source of infection, extent of infection dissemination [60] | Heterogenous infection in terms of bacterial pathogens and antibiotic resistance patterns, source of infection, e.g. catheter-associated versus dissemination from solid organ infections, immunocompromising conditions, dissemination to other organ systems<br>Various antibiotic regimens used, e.g. oral versus intravenous, various bioavailability in the bloodstream and organ systems |
| Gram-positive | 1 | Mortality;<br>microbiological eradication;<br>recurrence | > 2 weeks especially in *Staphylococcus aureus* bloodstream infections where there is low threshold to treat >4 weeks, based on expert opinion and observational data | Few trials evaluated antibiotic duration in staphylococcal bloodstream infections. The most recent trial enrolled a large proportion of patients who had coagulase-negative staphylococcal bloodstream infections [61]<br>Optimal management of *Staphylococcus aureus* bloodstream infections also involve infectious diseases consultation, thorough evaluation for infection source and secondary dissemination e.g. echocardiography, which have to be<br>optimised along with antibiotic duration [62] |
| **Others** |  |  |  |  |
| Bacterial meningitis, non-tuberculous | 7 | Clinical response; microbiological eradication (in CSF, in nasopharynx for *Neisseria meningitidis*); recurrence | Typically >10 days in adults, e.g. 10-14 days in Pneumococcal meningitis, >21 days in Gram-negative bacilli, based on expert opinion<br>Typically >10 days in neonates depending on CSF and blood cultures, clinical response and bacterial pathogen, e.g. 14 days for uncomplicated Group B Streptococcal meningitis, >21 days for Gram-negative enteric bacilli meningitis [63,64]<br>5 to 28 days depending on CSF and blood cultures, clinical response and bacterial pathogen, e.g. 5 to 7 days for *Neisseria meningitidis*, 21 to 28 days for *Listeria monocytogenes* | Mainly studied in neonates and children, no trials evaluated antibiotic duration in adults<br>Though a 2009 meta-analysis found that 4 to 7 days was similar to 7 to 14 in terms of clinical cure rate, it also highlighted important methodological issues including lack of blinding, small sample sizes, relatively short follow-up, high variability in the causative pathogens [65] |
| Skin and soft tissue infections (cellulitis and skin abscesses) | 10 | Clinical response; recurrence | 5 to 6 days for uncomplicated cellulitis depending on clinical response[66–69]<br>No additional benefit for systemic antibiotics for uncomplicated skin and soft tissue abscesses | No trials compared antibiotic duration for complicated soft tissue infections, e.g. recurrent cellulitis or abscesses<br>In the key trial which supported 5 days antibiotics for uncomplicated cellulitis, enrolled patients were only randomised<br>if clinical response was observed at day 5 of treatment. Definition of clinical response was poorly defined and was at the discretion of the treating physician [66] |
| Intra-abdominal infections | 7 | Clinical response; mortality; recurrence | 4 to 5 days given adequate surgical control of infection source [70,71]<br>4 to 6 weeks for pyogenic liver abscesses based on expert opinion | Lack of trials which compared antibiotic duration for intra-abdominal infections with inadequate source control, and pyogenic liver abscess<br>Heterogeneous infection in terms of infection sites, adequacy of source control, bacterial pathogens and antibiotic resistance patterns |
| Typhoid fever | 5 | Clinical response; microbiological eradication from gut; recurrence | 7 to 14 days or 5 days after fever resolution, whichever is longer, based on expert opinion [72]<br>Uncomplicated disease can be treated with 3 to 5 days of ceftriaxone and ofloxacin [73] | Few trials evaluated antibiotic treatment duration<br>Existing trials enrolled small numbers of patients, and compared various antibiotic regimens which makes direct comparison for treatment duration challenging |
| Post-surgical prophylaxis | 47 | Surgical site infection | Generally single dose or repeated dosing <24 hours is adequate, which is associated with comparable efficacy at preventing surgical site infections and less side effects [74–80] | Single dose or <24 hours of antibiotic prophylaxis is the general recommendation. However, gaps remain in defining redosing frequencies especially in certain specialised surgeries, e.g. cardiothoracic surgeries, where surgeries may be complicated and prolonged [81] |

The overall mortality reported in the trials was low (median 0%, IQR 0 to 2%). 62 trials (48%) reported the frequency of observed antibiotic side effects. 64 trials (50%) obtained follow-up samples from the participants to check for clearance or newly-acquired antibiotic resistant bacteria at the sites of infection. Only 9 trials (7%) took surveillance cultures during follow-up to assess for emergence of resistant bacteria colonisation, most commonly at upper respiratory, urinary or gastrointestinal tracts. 56 trials (44%) reported a health-economic outcome, the most common being length of hospital stay (47/56, 84%).

#### • Non-inferiority margin

About half of the trials used a non-inferiority design (68/128, 53%). Out of these, 61 reported a non-inferiority margin (90%) and all except one used absolute difference between the observed outcomes in the randomisation arms to calculate trial estimates. The mean non-inferiority margin was 10%, ranging from 0.35% to 25%. Figure 5 shows the observed proportion of trial participants who suffered negative clinical outcomes reported in these non-inferiority trials. The median proportion of participants having a negative clinical outcome was 13 (IQR 7% to 22%).

**Figure 5:**
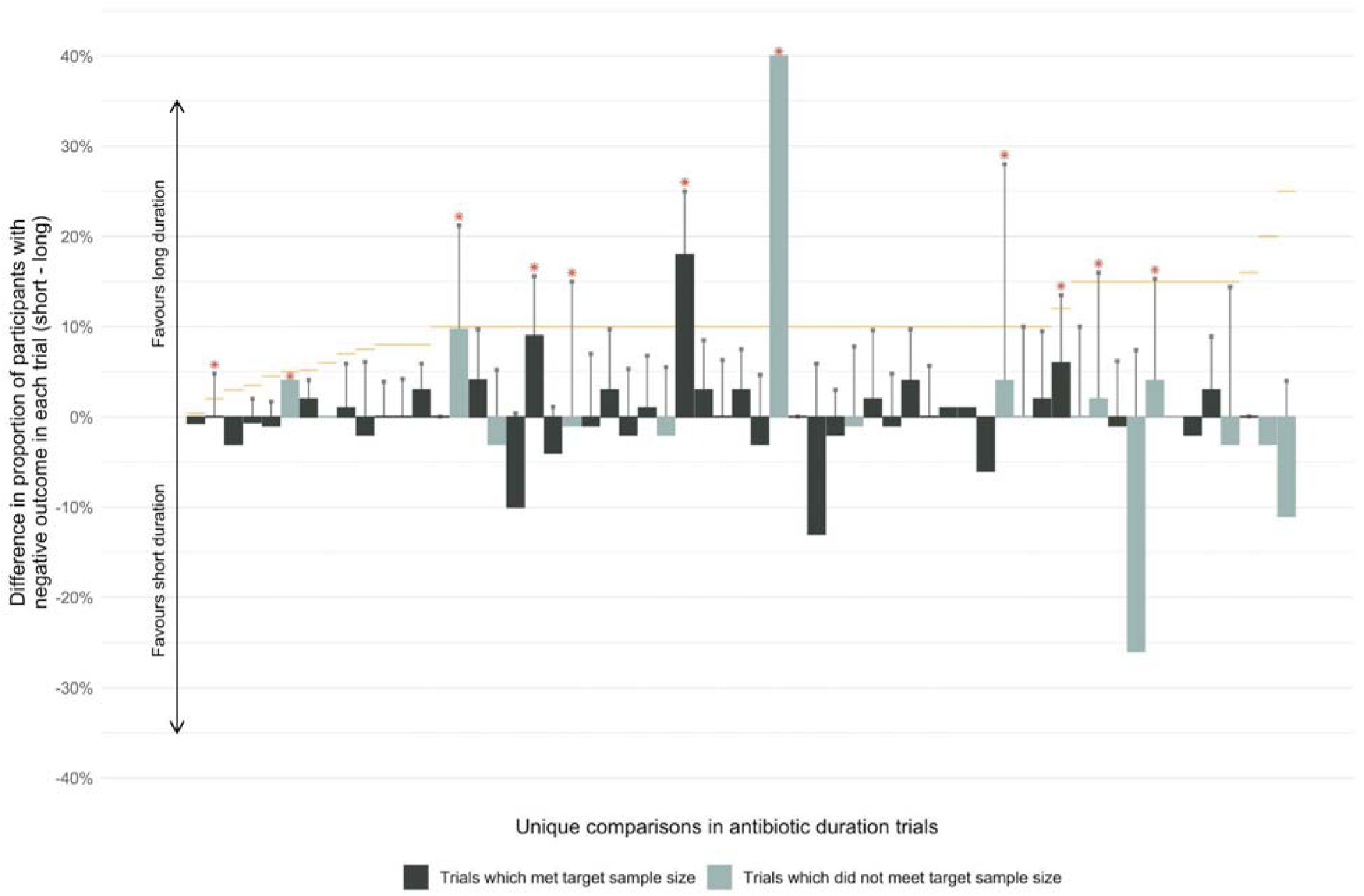
Trial estimates and respective non-inferiority margins in antibiotic duration non-inferiority trials. Each vertical bar represents a unique comparison between a long and a short duration. 59 non-inferiority trials which reported clinical outcomes as the primary outcome are shown. The bars represent the absolute difference in the proportion of participants with negative clinical outcome in each comparison (point estimates calculated by the proportion of participants with negative outcome in the short arm minus that in the long arm). The light coloured bars represent trial comparisons that did not meet the target sample size. Trial comparisons that failed to conclude non-inferiority are marked with a red asterisk. These are the trials in which the upper bound of the confidence intervals (indicated by grey vertical error bars for trials which reported the upper bounds) crossed the non-inferiority margins (indicated by yellow horizontal segments). None of the trials concluded that a short duration was superior to long duration based on the primary outcomes.

**Figure 6:**
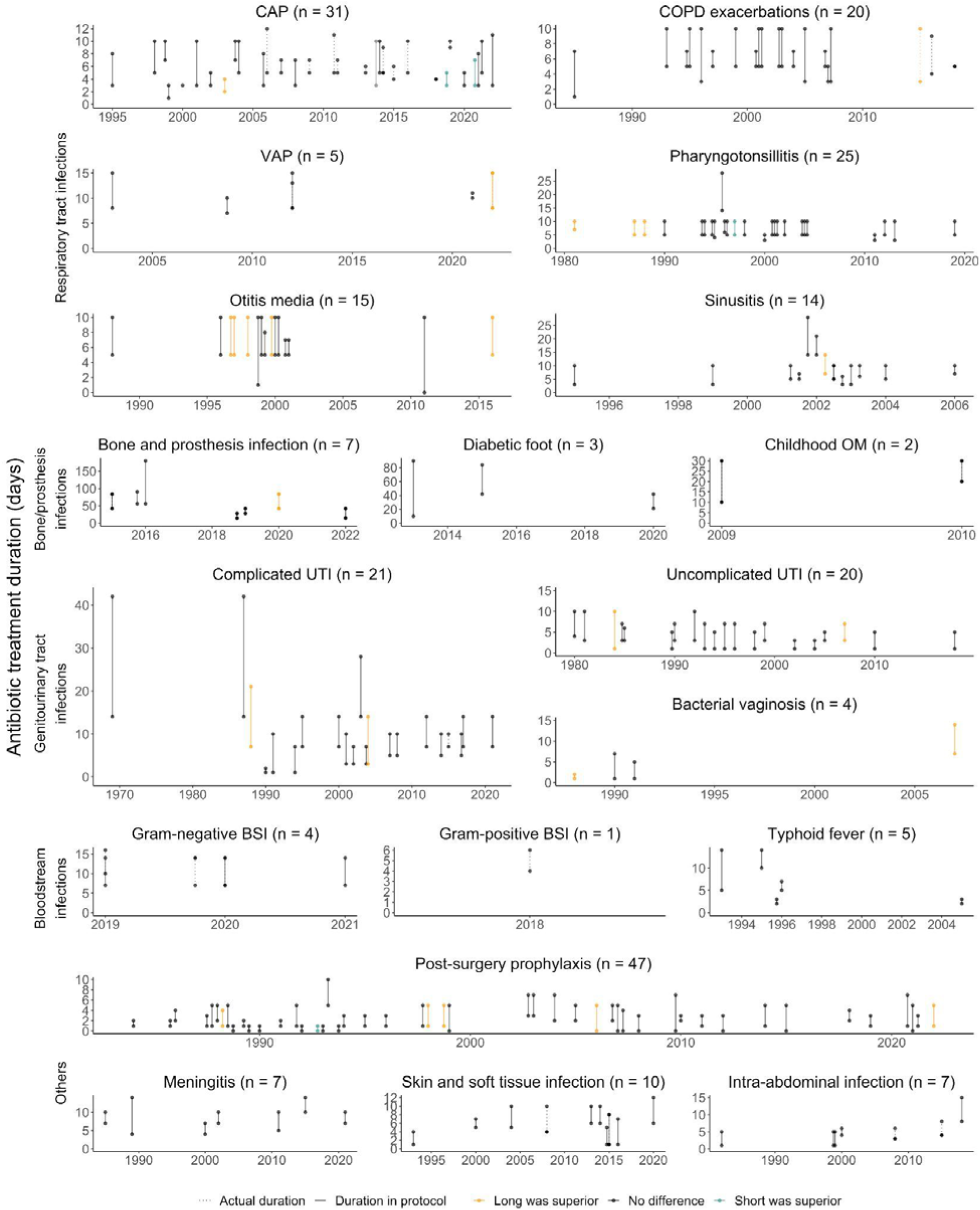
Antibiotic duration trials classified by bacterial infection syndromes. Each panel presents trial results for a type of bacterial infection. The number of trials included in each panel is shown in brackets. The vertical lines joining the points in each plot represent the durations compared in each trial. Line colours indicate whether trials concluded that long duration was superior to short duration (orange), not different or non-inferior (black), and inferior (teal) in terms of clinical outcomes. Solid vertical lines show the durations allocated to the study participants (plotted only when a trial defined arbitrary durations as the intervention). Dotted vertical lines show the actual duration observed during the trial (plotted only when reported). Abbreviations: CAP - community-acquired pneumonia, VAP - ventilator-associated pneumonia, COPD - chronic obstructive pulmonary disease, BSI - bloodstream infection, DM - diabetes mellitus, OM - osteomyelitis, UTI - urinary tract infection. Complicated UTI encompassed simple cystitis in males, catheter-associated UTI and pyelonephritis. The single trial shown in the Gram-positive BSI panel studied staphylococcal BSI. The typhoid fever panel includes duration trials for both complicated and uncomplicated typhoid fever. 4 trials in the systematic review were excluded from this figure as the antibiotic durations in these trials were vastly different from the other trials within the same infection syndrome. [82–85]

#### • Bias assessment

69 trials (69/128, 54%) reported sample size calculations and completed enrolment. Assessment of the trials with the RoB2 tool identified 25 (36%), 40 (58%), and 4 (6%) trials to be at high risk, with some concerns, and low risk of bias respectively (Supplementary material 2). The domain which led to most studies being classified as having high risk of bias was because many of these trials were unblinded, suffered from a high degree of non-adherence or crossovers, and that the analysis failed to account adequately for non-adherence. These factors could potentially bias the trial estimates.

## Discussion

In this systematic review of antibiotic duration trials published to date, most trials (85%, 253/296) successfully concluded that short-course treatment was comparable or non-inferior to longer courses. These trials mostly studied infections associated with low mortalities, such as respiratory tract and genitourinary infections, and post-surgical prophylaxis (73%, 215/296). Patients from low-to-middle-income countries, critically-ill patients and children were under- represented. Using arbitrarily-determined duration was the most common trial intervention throughout the years (90%, 266/296), while using biomarkers to stop antibiotics declined since the mid-2010s. Non-adherence to allocated antibiotic duration was prevalent (94%, 73/78 trials which reported per-protocol number of participants) and involved a median proportion of 13% (IQR 7 to 23%) in each trial. Few trials (7%, 9/128 trials published since 2006) took surveillance cultures to assess for resistant bacteria colonisation with various antibiotic duration.

Some of these trials were successful in changing stewardship policies for shortening treatment duration, such as in community-acquired pneumonia and uncomplicated urinary tract infections (Table 1). However, there are also conditions in which the practice guidelines have continued to support long treatment duration despite numerous randomised trials showing non-inferiority of a short course in terms of clinical outcomes. These include infections which require longer follow-ups (e.g. immunological sequelae of Group A Streptococcal infections), in low-to-middle-income countries where antimicrobial resistance to first-line empirical antibiotics is prevalent (e.g. otitis media in children), and highly heterogeneous infections for which trials with broad inclusion criteria were not informative but when inclusion criteria is too specific participant accruement becomes challenging (e.g. bacterial meningitis, prosthetic joint infections). There remain few randomised trials defining antibiotic duration for relatively common conditions such as Gram-negative and *Staphylococcus aureus* bloodstream infections and ventilator-associated pneumonia caused by non-fermenting Gram-negative bacilli.

Methodologically, a common source of potential bias arose due to the lack of blinding in antibiotic duration trials (69%). Treating physicians or patients, who were aware of infection severity, might become more careful during follow-up care or extend antibiotic duration regardless of randomisation. Likewise, unblinded assessors who determine the final outcomes in these trials might be influenced by their personal bias. Disease severity is therefore a potentially important confounder in both adherence to allocated duration and outcome assessment in antibiotic duration trials.

There is limited evidence to show the effectiveness of shortening antibiotic duration in reducing AMR. The effect of treatment duration on the development and spread of AMR is expected to vary due to multiple pathogens, host, and environmental factors. The penalties of prolonged antibiotic courses are the side effects and associated costs for both treated individuals and the population. Direct side effects of antibiotics for the treated individuals include allergies, kidney and liver injuries, and opportunistic infections such as *Clostridium difficile* colitis. Though mostly mild, these side effects are not infrequent and have been reported in up to 20% of hospitalised patients who received antibiotics. [86, 87] While there are sound theoretical reasons for expecting reduced treatment duration to often lead to reduced AMR, surprisingly little empirical evidence has been collected to explore this relationship. As noted in this review, the majority of antibiotic duration trials investigated the short-term clinical outcomes of reduced antibiotic duration on individual patients, but only 9 followed up on treated patients to determine subsequent AMR colonisation. This is echoed by earlier reviews of studies attempting to quantify the impact of stewardship strategies, which found that microbiology outcomes are seldom reported, especially those from longer term follow-ups. [6]

Non-inferiority was the most commonly adopted design for antibiotic duration trials. Non-inferiority trials compare the short with the standard treatment duration against a margin which the investigators are willing to sacrifice in terms of treatment effects, to reap benefits such as reduced resistance, cost savings or reduced side effects. A short treatment can be shown to be non-inferior if its effect is “no worse than” the standard duration by this margin of tolerance when compared to the standard duration. The choice of non-inferiority margin is frequently a subject of debate. [88–90] The non-inferiority margin is usually defined with prior knowledge of the standard-of-care antibiotic duration’s efficacy compared with a placebo, and consensus from subject matter experts. [91] However prior placebo-control trials to inform the choice of the non-inferiority margin are frequently lacking, and the efficacy of standard-of-care antibiotic duration may be increasing with time due to improvements in general healthcare delivery. [92] In addition, the actual primary outcome event rate in the trial should also be monitored and compared against that used in the power calculations. The systematic review showed that 90% of the antibiotic duration non-inferiority trials published in the last 15 years reported a very low event rate of <10%. A lower than expected event rate can lead to insufficient power, favouring non-inferiority. [93]

Another caveat for interpreting non-inferiority trials is that features concerning disease definition, treatment, and outcome assessment may result in outcomes in the randomisation groups appearing more similar, which will increase the probability of demonstrating non-inferiority even when the short duration is actually inferior in clinical efficacy.[94] In addition, lapses in the conduct of the trial, such as non-adherence, are potentially not generalisable, and should be minimised. Though commonly reported, non-adherence in non-inferiority trials is not adequately addressed in terms of analysis methods and reporting from international guidelines. [88, 91]

Reducing treatment duration is likely to remain an important strategy for antibiotic stewardship, and an area of active research. Innovative and robust trial designs have been proposed to define duration-response relationships, [95] rather than relying on arbitrary durations compared with non-inferiority hypotheses. Larger randomised trials will allow for meaningful conclusions for patient subsets with varying host and pathogen characteristics. Areas which require more evidence to define treatment duration include in severe bacterial infections and low-to-middle-income settings.

## Data Availability

All data produced are available online at https://github.com/moyinNUHS/abxduration_abm.

## Contributors

MY conceptualised and drafted the first version of this manuscript. MY and TWC performed the systematic review independently and cross-checked data extracted. All authors reviewed and edited the manuscript.

## Data Sharing

Data extracted from all included antibiotic treatment duration randomised controlled trials are available at https://github.com/moyinNUHS/abxduration_abm.

## Acknowledgements

We acknowledge Professor Sarah Walker and Professor Marc Bonten for their helpful insights in refining this manuscript.

## Supplementary material 1

**Table S1:** Search terms used in the literature review.

| Database | Search Term |
| --- | --- |
| MEDLINE | ((antibiotic) AND (infection)) AND (weeks[Title])<br>**Filters:** Randomized Controlled Trial, Humans, 1920–2021<br>**Details:** (((((((("anti bacterial agents"[Pharmacological Action] OR "anti-bacterial agents"[MeSH Terms]) OR ("anti bacterial"[All Fields] AND "agents"[All Fields])) OR "anti bacterial agents"[All Fields]) OR "antibiotic"[All Fields]) OR "antibiotics"[All Fields]) OR antibiotics"[All Fields]) OR "antibiotical"[All Fields]) AND (((((((((((((((("infect" [All Fields] OR "infectability" [All Fields]) OR "infectable"[All Fields]) OR "infectant"[All Fields]) OR "infectants"[All Fields]) OR "infected"[All Fields]) OR "infecteds" [All Fields]) OR "infectibility"[All Fields]) OR "infectible" [All Fields]) OR "infecting"[All Fields]) OR "infections" [All Fields]) OR "infections"[MeSH Terms]) OR "infections"[All Fields]) OR "infection"[All Fields]) OR "infective"[All Fields]) OR "infectiveness" [All Fields]) OR "infectives"[All Fields]) OR "infectivities"[All Fields]) OR "infects" [All Fields]) OR "pathogenicity"[MeSH Subheading]) OR "pathogenicity"[All Fields]) OR "infectivity" [All Fields])) AND "weeks"[Title] |
|  | ((antibiotic) AND (infection)) AND (days[Title])<br>**Filters:** Randomized Controlled Trial, Humans, 1920–2021<br>**Details:** (((((((("anti bacterial agents"[Pharmacological Action] OR "anti-bacterial agents"[MeSH Terms]) OR ("anti bacterial"[All Fields] AND "agents"[All Fields])) OR "anti bacterial agents"[All Fields]) OR "antibiotic"[All Fields]) OR "antibiotics"[All Fields]) OR antibiotics"[All Fields]) OR "antibiotical"[All Fields]) AND (((((((((((((((("infect" [All Fields] OR "infectability" [All Fields]) OR "infectable"[All Fields]) OR "infectant"[All Fields]) OR "infectants"[All Fields]) OR "infected"[All Fields]) OR "infecteds" [All Fields]) OR "infectibility"[All Fields]) OR "infectible" [All Fields]) OR "infecting"[All |
|  | Fields]) OR "infections" [All Fields]) OR "infections"[MeSH Terms]) OR "infections"[All Fields]) OR "infection"[All Fields]) OR "infective"[All Fields]) OR "infectiveness" [All Fields]) OR "infectives"[All Fields]) OR "infectivities"[All Fields]) OR "infects"[All Fields]) OR "pathogenicity"[MeSH Subheading]) OR "pathogenicity"[All Fields]) OR "infectivity" [All Fields])) AND "days"[Title]<br>((antibiotic) AND (infection)) AND (duration)<br>**Filters:** Randomized Controlled Trial, Humans, 1920–2021<br>**Details:** (((((((("anti bacterial agents"[Pharmacological Action] OR "anti-bacterial agents"[MeSH Terms]) OR ("anti bacterial"[All Fields] AND "agents"[All Fields])) OR "anti bacterial agents"[All Fields]) OR "antibiotic"[All Fields]) OR "antibiotics"[All Fields]) OR "antibiotic s"[All Fields]) OR "antibiotical"[All Fields]) AND (((((((((((((((("infect"[All Fields] OR "infectability"[All Fields]) OR "infectable"[All Fields]) OR "infectant"[All Fields]) OR "infectants"[All Fields]) OR "infected"[All Fields]) OR "infecteds" [All Fields]) OR "infectibility"[All Fields]) OR "infectible"[All Fields]) OR "infecting"[All Fields]) OR "infections"[All Fields]) OR "infections"[MeSH Terms]) OR "infections"[All Fields]) OR "infection"[All Fields]) OR "infective"[All Fields]) OR "infectiveness"[All Fields]) OR "infectives"[All Fields]) OR "infectivities"[All Fields]) OR "infects"[All Fields]) OR "pathogenicity"[MeSH Subheading]) OR "pathogenicity"[All Fields]) OR "infectivity"[All Fields])) AND ("duration"[All Fields] OR "durations" [All Fields])) |
| EMBASE | antibiotic AND infection AND week*:ti AND [randomized controlled trial]/lim AND [1920–2021]/py<br>AND 'human'/de<br>antibiotic AND infection AND day*:ti AND [randomized controlled trial]/lim<br>AND [1920–2021]/py<br>AND 'human'/de<br>antibiotic AND infection AND duration AND [randomized controlled trial]/lim AND [1920–2021]/py<br>AND 'human'/de |

## Supplementary material 2

### Risk of Bias Tool 2 assessment

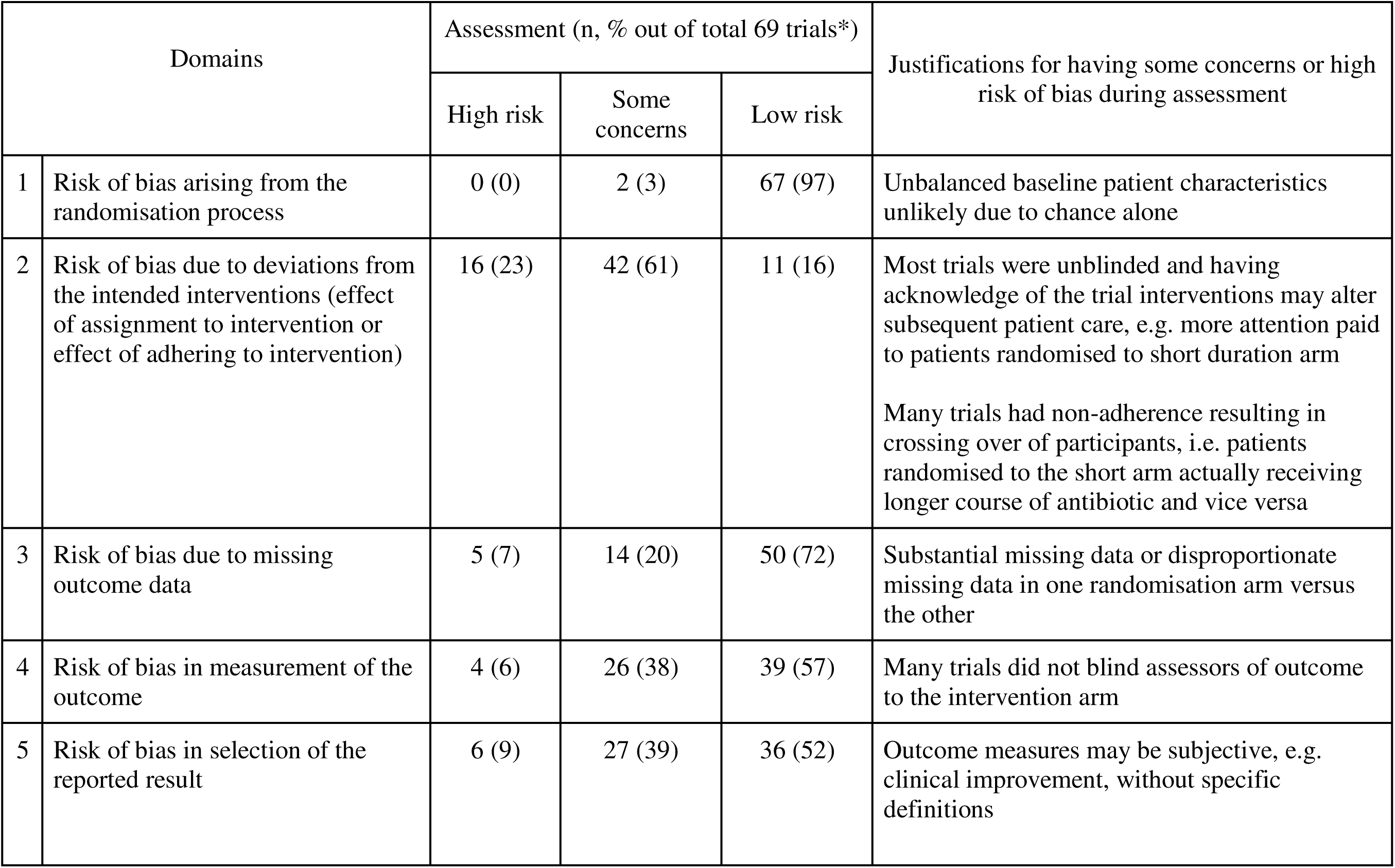
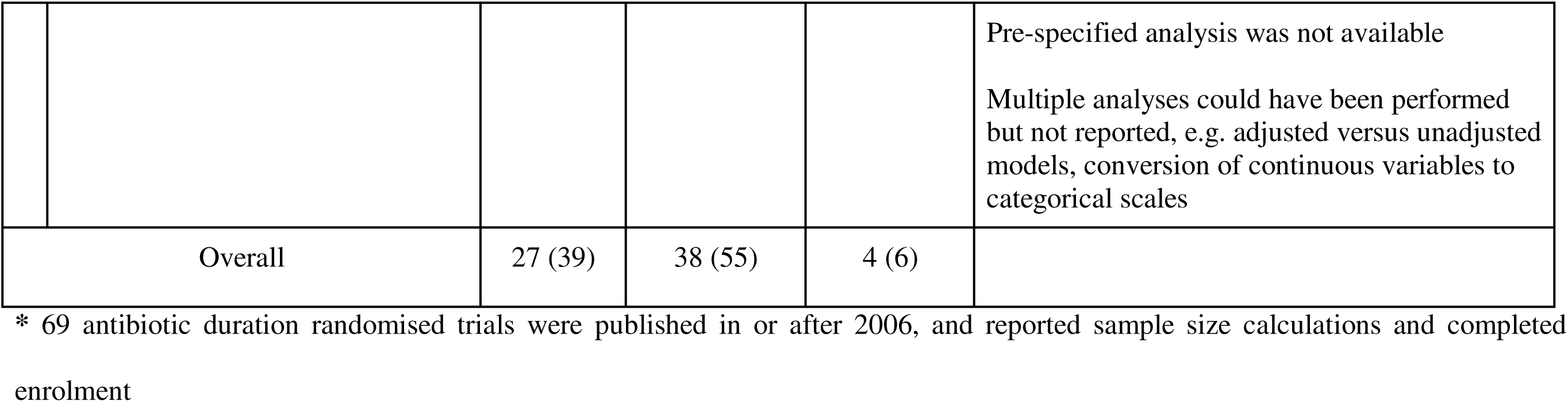

Supplementary material 2 Risk of Bias Tool 2 assessment
| Domains |  | Assessment (n, % out of total 69 trials*) |  |  | Justifications for having some concerns or high risk of bias during assessment |
| --- | --- | --- | --- | --- | --- |
|  |  | High risk | Some concerns | Low risk |  |
| 1 | Risk of bias arising from the randomisation process | 0 (0) | 2 (3) | 67 (97) | Unbalanced baseline patient characteristics unlikely due to chance alone |
| 2 | Risk of bias due to deviations from the intended interventions (effect of assignment to intervention or effect of adhering to intervention) | 16 (23) | 42 (61) | 11 (16) | Most trials were unblinded and having acknowledge of the trial interventions may alter subsequent patient care, e.g. more attention paid to patients randomised to short duration arm<br>Many trials had non-adherence resulting in crossing over of participants, i.e. patients randomised to the short arm actually receiving longer course of antibiotic and vice versa |
| 3 | Risk of bias due to missing outcome data | 5 (7) | 14 (20) | 50 (72) | Substantial missing data or disproportionate missing data in one randomisation arm versus the other |
| 4 | Risk of bias in measurement of the outcome | 4 (6) | 26 (38) | 39 (57) | Many trials did not blind assessors of outcome to the intervention arm |
| 5 | Risk of bias in selection of the reported result | 6 (9) | 27 (39) | 36 (52) | Outcome measures may be subjective, e.g. clinical improvement, without specific definitions |
|  |  |  |  |  | Pre-specified analysis was not available<br>Multiple analyses could have been performed but not reported, e.g. adjusted versus unadjusted models, conversion of continuous variables to categorical scales |
|  | Overall | 27 (39) | 38 (55) | 4 (6) |  |

\* 69 antibiotic duration randomised trials were published in or after 2006, and reported sample size calculations and completed enrolment

